# Genotoxicity of Prenatal and Early Childhood Exposure to Pesticides: A Protocol and Pilot Study of a Systematic Review and Meta-Analysis

**DOI:** 10.1101/2024.08.29.24312778

**Authors:** Moustafa Sherif, Aya Darwish, Balázs Ádám

## Abstract

1.

**Objectives:** To systematically review and meta-analyse the genotoxic impact of prenatal and early childhood pesticide exposure, investigating prevalence, specific pesticides, effect size, mechanisms, genetic susceptibility, and vulnerable periods.

**Study Design:** A protocol for systematic review and meta-analysis. A pilot study was also conducted to develop appropriate extraction and risk of bias tool.

**Methods:** Adhering to 2020 PRISMA guidelines, the review will explore genotoxic impact of prenatal and early childhood pesticide exposure in children up to 5 years. The protocol had been registered in the International Prospective Register of Systematic Reviews (PROSPERO) database (CRD42024510877). Searches was done across PubMed, EMBASE, Web of Science, and Scopus use keywords ‘prenatal or childhood’, ‘pesticides’, and ‘genotoxicity’. Manual reference screening supplements searches. Eligible observational studies (cross-sectional, case-control, cohort designs) in English will be included, while excluding case reports and in vitro studies, using Covidence screening tool. Two independent reviewers will use Newcastle-Ottawa Scale and novel tool for cross-sectional studies for screening, data extraction, and risk of bias assessment. Findings will be synthesized narratively, with potential meta-analysis of genotoxicity outcomes. GRADE approach will assess the evidence quality.

**Results:** A pilot test screened 1,405 studies, resulting in 21 eligible for full-text screening. Twelve were excluded. Data extraction and risk of bias assessment followed pre-defined protocols. Findings informed the refinement of study procedures.

**Conclusions:** The protocol outlines a comprehensive approach to systematically review the genotoxic impact of prenatal and early childhood pesticide exposure, aiming to provide necessary insights to a better understanding of this environmental risk.

## 2. Introduction

Extensive pesticide use has resulted in widespread exposure that raised human health concerns ^1,2^. Vulnerable populations, especially pregnant women and children, face ongoing environmental pesticide exposure due to inadequate knowledge among fieldworkers, lax regulatory control, and economic interests. The idea behind the Developmental Origin of Human Disease (DOHAD) theory, is that the conditions we experience in the early stages of life can affect our health later on ^3^.

The International Agency for Research on Cancer (IARC) categorizes certain pesticides as either carcinogenic or potentially carcinogenic for humans ^4^. This classification takes into consideration their ability to induce DNA damage and/or mutations in somatic or gametic cells. Scientific evidence supports the hypothesis that pesticide exposure by itself or with other environmental agents, is a significant risk factor for serious diseases, including reproductive disorders, neurological and metabolic alterations, particularly in exposed children ^5^. Epidemiological studies provide substantial evidence linking pesticide exposure with persistent and biomodulating properties, such as oganochloride compounds (OCs), to pediatric leukemia, constituting 30% of all childhood cancers ^5,6^.

There are several biomarkers indicating genotoxic damage and genomic instability in cells, such as chromosomal aberrations (CA) and micronuclei (MN) ^7,8^. If the DNA damage is not naturally repaired by cellular DNA repair enzyme mechanisms or if the affected cell is not eliminated, there is a risk that the flawed cell experiences changes in its physiological or metabolic functions ^9^. This form of damage is recognized as one of the primary mechanisms underlying chronic diseases, particularly in the contexts of carcinogenesis and teratogenesis. Genotoxicological biomonitoring is viewed as an essential component of thorough medical monitoring for individuals exposed to pesticides ^7^. This method enables the assessment of potential risks associated with exposure to genotoxic compounds in the environment, such as pesticides. Additionally, it facilitates the early detection of alterations, contributing to the prevention of chronic diseases, such as cancer.

Our overarching goal is to systematically review the scientific literature and conduct a meta-analysis on the genotoxic impact of prenatal and early childhood exposure to pesticides. To achieve this goal, our objectives include identifying and evaluating research articles to synthesize information on the prevalence, sources, and routes of pesticide exposure in the prenatal and early childhood period with genotoxic effects. Additionally, we aim to quantify the size of the effect, the strength of association between pesticide exposure and genotoxicity, identifying genotoxic pesticides, exploring mechanisms, and assessing susceptibility factors, including potential vulnerable periods during prenatal and early childhood stages.

## 3. Methods/Design

The systematic review will follow the standardized methodology outlined in the 2020 Preferred Reporting Items for Systematic Reviews and Meta-analyses (PRISMA) statement, which incorporates the Grading of Recommendations Assessment, Development and Evaluation (GRADE) approach for environmental health assessments ^11,12^. This protocol aligns with the Preferred Reporting Items for Systematic Reviews and Meta-analyses Protocol (PRISMA-P) statement ^13^. The modified PRISMA-P checklist for Environment International is available in supplementary file S1. The protocol had been registered in the International Prospective Register of Systematic Reviews (PROSPERO) database (CRD42024510877).

### Eligibility criteria

To assess the genotoxic effects of prenatal and early childhood pesticide exposures, a Population, Exposure, Comparator, and Outcomes (PECO) statement was formulated as the following ^14^.

#### A. Participants/population

Inclusion: Embryos, fetuses, infants and young children up to 5 years of age. Pregnant mothers will be considered if the placenta was investigated.

Exclusion: Children older than 5 years of age, non-human subjects or cells in toxicological and mechanistic studies, and pregnant mothers who manifest genotoxicity in tissues other than the placenta.

#### B. Intervention(s), exposure(s)

Inclusion: The study will consider exposure to pesticides during the prenatal and/or early childhood periods below 5 years of age, irrespective of specific details on the types of pesticides, their metabolites, or the route of exposure. Additionally, exposures evidenced by biomarkers of pesticide exposure in mothers will be included, drawing associations with the level of genotoxicity in their paired children. Exposure through maternal blood, placental transfer, or breastfeeding will also be considered.

Exclusion: The study will exclude individuals exposed to environmental factors other than pesticides and those exposed to pesticides during time periods other than the prenatal and/or early childhood below 5 years of age.

#### C. Comparator/Control

A comparison group (e.g., unexposed or low-exposure group) is essential for evaluating the association/effect size. However, for research focused on exposure prevalence and the mechanisms of effects, no comparator group is necessary. Mothers can serve as comparators for their paired infants.

#### D. Outcome

Inclusion: The systematic review will incorporate studies focusing on the genotoxic consequences of prenatal and early childhood pesticide exposure. Eligible outcomes encompass genetic damages measured through biomarkers of genotoxicity assays (e.g., comet assay, micronucleus test, chromosomal aberrations), associations between prenatal/early childhood pesticide exposure and genotoxicity, and the role of genetic polymorphisms in heightened susceptibility to genotoxicity induced by pesticide exposure in the study population.

Exclusion: Studies reporting outcomes beyond the predefined categories, such as allergies, infectious diseases, and childhood cancers (including leukemia and lymphoma), without associated genetic damages or mutations will be excluded. This ensures a focused investigation specifically on the impact of pesticide exposure on genotoxicity.

### Types of studies

The review will include observational studies, such as cross-sectional, case-control, and cohort studies to address the research objectives. Conversely, excluded study types encompass *in vitro* and *in vivo* animal studies, case reports, opinion articles, commentaries, letters, review articles, published abstracts, and conference proceedings. Adhering to these criteria ensures a focus on high-quality original studies that directly contribute to the research objectives.

### Search strategy

In February 2024, we conducted a pilot systematic search using electronic databases, namely PubMed (NLM), EMBASE (Elsevier), Web of Science (Clarivate), and Scopus (Elsevier), to identify relevant studies. The search strategy was based on the pre-defined PECO statement and adhered to the PRESS Peer Review of Electronic Search Strategies ^15^. MS conducted the search in accordance with the PRISMA-S extension ^16^. The initial search strategy was developed in PubMed and systematically adapted and replicated across all selected databases.

The search involves applying a combination of search fields, including “title,” “abstract,” and PubMed’s Medical Subject Headings (MeSH) terms (for PubMed), with no restrictions on publication year or language as displayed in Fig. 1A. Keywords with synonyms for search terms revolving around key concepts was performed as illustrated in Fig.1B. The search domains include ‘prenatal’ for the period before birth during pregnancy, or ‘childhood’ for the period following birth, specifically targeting the first five years of a child’s life to explore potential genotoxicity resulting from early pesticide exposure. The second domain, linked by “AND” includes terms related to pesticides, while the third domain, also connected by “AND” encompasses terminology associated with genotoxicity. The search string created in pilot searches will be re-executed before the conclusive analysis. The most recent reproducible search strings as of February 2024, along with results and technical notes, are available in supplementary file S2. The comprehensive search logs, encompassing search terms, results, and technical notes (including search date and any adjustments), for all databases will be included in the review.

**Fig. 1.**
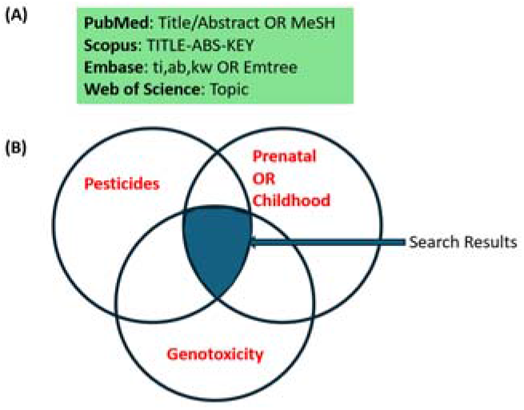
Search fields per each of the databases (A), and a graphical representation of the conceptual search domains utilized in formulating the search terms for electronic databases (B).

### Study records

#### A. Selection process

The search outcomes will be imported into the systematic review tool of Covidence ^17^, where the screening process will be documented. After eliminating duplicates, two independent reviewers (MS and AD) will assess the titles and abstracts of unique studies against inclusion and exclusion criteria. Subsequently, full-text screening of the included studies will be performed by the same two experienced reviewers (MS and AD). Manual screening through backward snowballing will involve examining the reference lists of all eligible reviewed articles and previously found systematic reviews to identify additional papers meeting the inclusion and exclusion criteria. In cases a large number of articles are identified (exceeding 150 papers), an alternative approach may involve focusing on relevant and highly cited articles. In instances of uncertainty, a third reviewer (BA) will determine the final decision. The screening and selection will be visually depicted in a PRISMA flow-diagram, similar to Fig. 2. To maintain transparency in the full-text screening process, justifications for exclusions, such as incorrect study design, population, exposure, comparator, or outcome, will be thoroughly reported. Finally, Cabell’s Predatory Reports will be utilized to verify the academic quality of eligible studies published in open access journals.

**Fig. 2.**
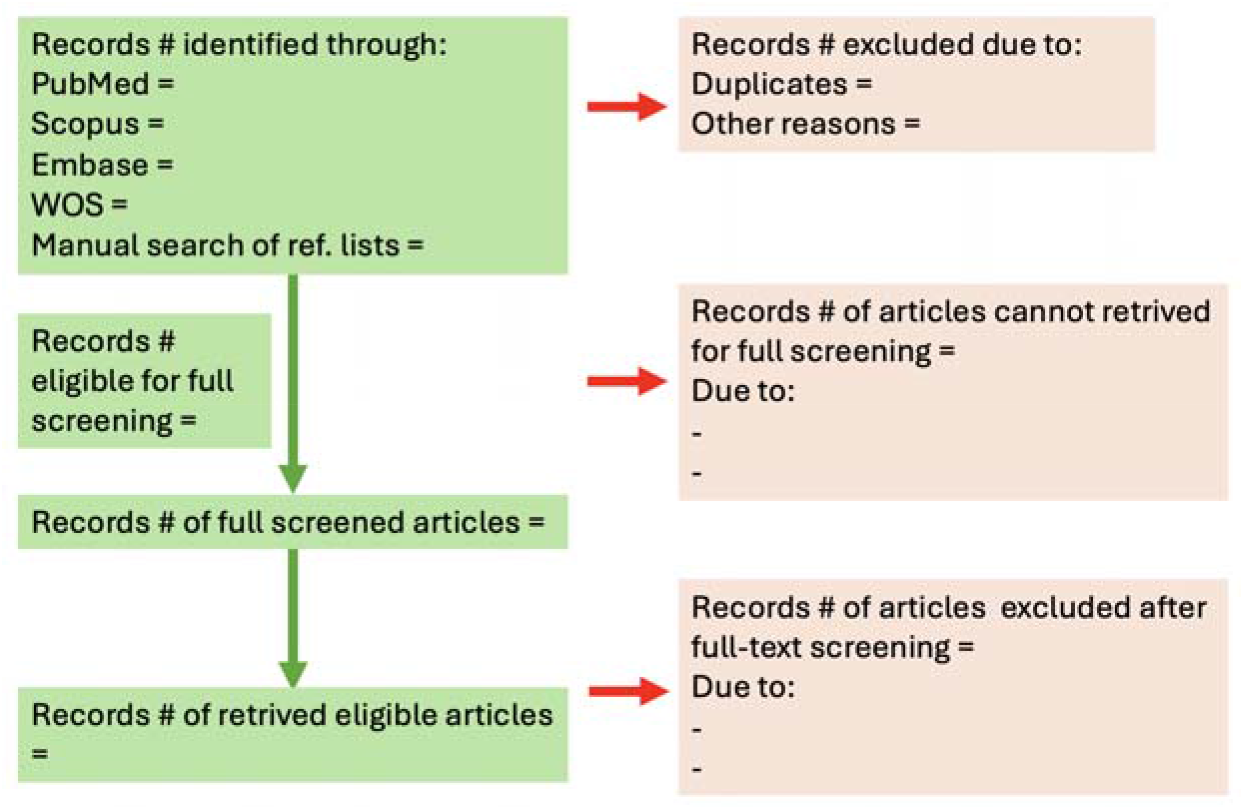
Example of the PRISMA flowchart to be employed in this study.

#### B. Data collection process

We created the data extraction template in Excel 2019 and tested it using nine articles (available in supplementary file S3). MS and AD will independently extracting data in this excel tool. Any discrepancies will be resolved through discussion with BA. We may reach out to study authors for any missing data. In cases when authors are unresponsive after two trials over two weeks or provide unclear data, the unavailable study results will be excluded from the review.

#### C. Data items

The following items will be extracted from the studies:

- Study identification (title, name of the first author, year of publication, DOI);
- Conflict of interest and ethical approval;
- Study characteristics (type of study, objectives, country, start and end date, response rate);
- Characteristics of exposed participants (number of children/infants, number of males, age, ethnicity, sources of recruitment, age of mother, occupation of the mother, proportion of previous abortions, proportion of diseased participants);
- Characteristics of reference group/comparator (identification of the reference group, number of participants, number of children/infant, number of males, age of children/infants, sources of recruitment, significant differences in characteristics with the exposure group);
- Exposure characteristics and exposure assessment (way of exposure, exposure assessment method, prevalence of exposure, duration of exposure, level of exposure);
- Outcomes
- DNA damage assessment (type of sample, assay, and DNA biomarker, plus number of samples/individuals, and units)
- Difference in level of exposure between exposed and reference groups (size, direction, level of significance);
- DNA damage (frequency, extent, direction of change, measure of association, statistical tests, significance);
- Confounders (confounders, adjustments)
- Correlation of DNA damage between mothers and their children (direction, size, significance).

### Risk of bias (RoB) assessment

Two skilled reviewers (MS and AD) will assess the risk of bias in the included studies using the Newcastle-Ottawa Scale (NOS) for case-control and cohort studies ^18^. Additionally, we have developed a novel risk of bias tool for evaluating RoB in cross-sectional studies, built upon the NOS framework. As illustrated in Figure 3, NOS is tailored to specific study types, evaluating three primary domains: selection, comparability, and exposure and/or outcome. The risk of bias for each domain will be determined by answering 1 to 4 RoB questions. A customized grade will be assigned to each domain, with classifications ranging from Definitely Low Risk of Bias to Definitely High Risk of Bias. The overall assessment will be influenced by the highest perceived risk in any of the individual bias domains. If uncertainties arise regarding the risk of bias in specific studies, a third reviewer (BA) will be consulted for resolution.

**Fig. 3.**
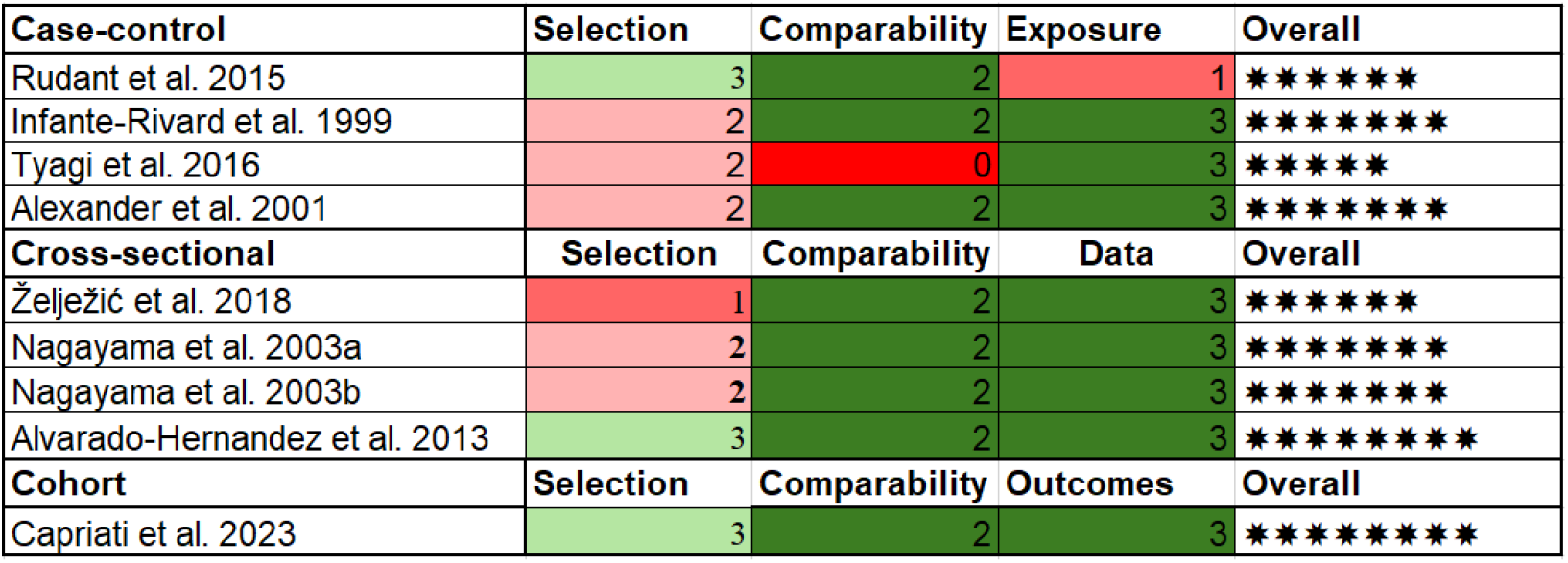
Results of the risk of bias assessment for the nine piloted studies presented using a study-specific Newcastle-Ottawa Scale (NOS) tool. The overall numbers of stars are specific to study type, and not comparable between different types of studies, with maximum numbers of 9 stars for all studies. Criteria for assessment are available in supplementary file S4.

### Data synthesis

The study findings will be initially described and synthesized narratively in accordance with the Synthesis Without Meta-Analysis (SWiM) reporting guideline ^19^. We will conduct a comprehensive narrative synthesis of findings from the included studies that will be tabulated by study-type, and organized by the type of pesticides investigated, population characteristics, and the nature of genotoxicity. This synthesis aims to offer a systematic overview of results and conclusions derived from primary research.

If two or more studies provide suitable estimates on outcome frequency and/or effect size, two reviewers will independently assess the clinical heterogeneity of the studies for potential meta-analysis. It is crucial to emphasize that the meta-analysis may specifically aim to quantify the effect size of genotoxicity resulting from prenatal/early childhood pesticide exposure in children. This quantitative analysis, in addition to the narrative synthesis, will offer a more precise evaluation of the strength and direction of the association between pesticide exposure and genotoxicity. The decision to conduct a meta-analysis will hinge on the availability and quality of data from selected studies, detailed within the systematic review.

If the combined studies exhibit sufficient clinical homogeneity, frequency and/or effect estimates will be pooled in a quantitative meta-analysis using the inverse variance method with a random-effects model to account for cross-study heterogeneity ^20^. Statistical heterogeneity will be analyzed using I^2^ statistics ^21^, and the meta-analysis will be conducted using RevMan software ^22^. Forest plots will illustrate the combined estimations. Non-quantitative and skewed data will be presented descriptively.

To visually assess publication bias, funnel plot graphics will be employed, and a sensitivity analysis will be conducted in the presence of outliers or asymmetry in the funnel plot.

### Subgroup and sensitivity analyses

Subgroup analysis and meta-regression will be performed to explore potential variations in the strength of association and effect size of genotoxicity estimates based on population characteristics, including country, prenatal/postnatal exposure, sex, type of pesticides or their specific metabolites, and the nature of genotoxicity. To evaluate the impact of individual studies on the pooled results, a sensitivity analysis will be conducted. This involves systematically excluding the effect estimate of each study one by one, considering both the RoB level, and recalculating the combined estimates based on the remaining studies. These analyses are crucial for a more in-depth exploration of potential differences within specific subgroups, offering valuable insights into the connection between pesticide exposure and its influence on genotoxicity in children.

### Quality of cumulative evidence (QoE)

Certainty of the evidence will be assessed by all reviewers using GRADE to assess individual RoB, inconsistency, indirectness, imprecision, and publication bias. GRADE scoring will allow us to provide summary-based evidence statements ^23^. The certainty of the estimated effect will be categorized into four levels ^24^: very low certainty of evidence (the true effect is likely to substantially differ from the effect estimate), low certainty of evidence (the true effect may significantly differ from the effect estimate), moderate certainty of evidence (the true effect is likely to be close to the effect estimate, but there is a possibility of substantial difference), and high certainty of evidence (the true effect aligns closely with the effect estimate).

## 3. Results

### Pilot test

The screening, data collection, and risk of bias (RoB) assessment processes were piloted using a random sample of studies (1,405 out of 6,945 unique records) identified through an extended pilot search across the pre-defined four electronic databases. Pilot results, uploaded and de-duplicated in Covidence, underwent screening by two reviewers (MS and AD) during title/abstract screening, resulting in the exclusion of 1,384 out of 1,405 studies. Twenty-one studies remained eligible for full-text screening. During full-text screening, twelve studies were excluded based on pre-defined criteria. Data extraction from the remaining nine studies adhered to the review protocol and was independently conducted by two reviewers (MS and AD). Subsequently, RoB assessment for the eligible studies was completed by the three main domains of the NOS RoB tool, with a risk of bias grade assigned for each domain of each studies. Supplementary Material 3, and 4 provide details on the pilot study data extraction, and RoB assessment, respectively. The overall risk of bias rating for the studies identified in the pilot test is presented in Figure 3.

## 4. Discussion

There is an increasing concern regarding the impact of pesticide applications on vulnerable populations such as pregnant women and children. This systematic review will assess the genotoxic consequences resulting from prenatal and early childhood exposure to pesticides, considering a range of outcomes, exposure sources, and potential susceptibility factors.

This systematic review has certain limitations, such as the potential for publication bias and language bias. The exclusion of certain study types, such as in vitro and in vivo animal studies, may limit a comprehensive understanding of the mechanisms underlying genotoxic effects. Moreover, the potential for heterogeneity among the included studies, arising from differences in populations, pesticides studied, and genotoxicity assessment methods, may impact the generalizability of the findings.

Despite these limitations, the study’s findings have the potential to inform preventive measures, public health policies, and interventions aimed at protecting the health and well-being of children.

## Data Availability

All data produced in the present work are contained in the manuscript.

## List of abbreviations

GRADE: Grading of Recommendations Assessment, Development and Evaluation
NOS: Newcastle-Ottawa Scale
PRISMA: Preferred Reporting Items for Systematic Reviews and Meta-analyses.
PECO: Population, Exposure, Comparator and Outcomes.
RoB: Risk of Bias.
QoE: Quality of cumulative Evidence

## Declarations

### Ethics approval and consent to participate

Ethics approval is not necessary for such type of research (systematic review), because no public members or patients will participate in the study. No patient consent is required for this study.

### Consent for publication

Not applicable

### Availability of data and materials

Data sharing is not applicable to this article as no datasets were generated or analysed during the current study.

### Competing interests

The authors declare that they have no known competing financial interests or personal relationships that could have appeared to influence the work reported in this paper.

### Funding

The contribution of MS was supported by the United Arab Emirates University PhD grant; while the work of AD was supported by the Pathfinder Epidemiology Academy [24PFA04, 2024].

### Authors’ contributions

Conceptualization and protocol development were undertaken by MS, AD, and BÁ. The search strategy was formulated by MS and AD. Literature search responsibilities will be carried out by MS and AD. Screening processes will be executed by MS and AD, and any conflicts will be resolved by BÁ. Data extraction will be conducted by MS and AD, with validation overseen by BÁ. MS and AD will perform the RoB assessment, and BÁ will resolve conflicts. The evaluation of the quality of evidence will involve all reviewers.

Acknowledgements

Not applicable

### Appendix. Supplementary material

PRISMA-P checklist: supplementary file S1

Pre-search string: supplementary file S2

Data extraction tool: supplementary file S3

Newcastle-Ottawa Scale (NOS) Risk of Bias (RoB) tool: supplementary file S4

## Notes

### Competing Interest Statement

The authors have declared no competing interest.

