## Supplementary file S2 for "Genotoxicity of Prenatal and Early Childhood Exposure to Pesticides: A Protocol and Pilot Study of a Systematic Review and Meta-Analysis"

| **Database** | **Search Terms** | **Hits and date** |
| --- | --- | --- |
| PubMed | ((toxicogenetic*[Title/Abstract] OR "Toxicogenetics"[MeSH] OR genotox*[Title/Abstract] OR "Mutagenicity Tests"[MeSH] OR "Carcinogenicity Tests"[MeSH] OR pharmacogenomic*[Title/Abstract] OR "Pharmacogenomic Testing"[MeSH] OR "Comet Assay"[MeSH] OR "comet assay*"[Title/Abstract] OR "Micronucleus Tests"[MeSH] OR "micronucleus test*"[Title/Abstract] OR "chromosome aberrat*"[Title/Abstract] OR "chromosomal aberrat*"[Title/Abstract] OR "Chromosome Aberrations"[MeSH] OR denaturation*[Title/Abstract] OR "Nucleic Acid Denaturation"[MeSH] OR "fluorescent in situ hybridization*"[Title/Abstract] OR "In Situ Hybridization, Fluorescence"[MeSH] OR "Nucleic Acid Hybridization"[MeSH] OR "nucleic acid hybridization*"[Title/Abstract] OR "ames test*"[Title/Abstract] OR aneuploidy*[Title/Abstract] OR "Aneuploidy"[Mesh] OR topoisomerase*[Title/Abstract] OR "DNA Topoisomerases"[Mesh] OR "Teniposide"[Mesh] OR teniposide*[Title/Abstract] OR "Etoposide"[Mesh] OR etoposide*[Title/Abstract] OR greenscreen*[Title/Abstract] OR "γH2AX*"[Title/Abstract] OR "pH2AX*"[Title/Abstract] OR "high content screening*"[Title/Abstract] OR "cycle arrest*"[Title/Abstract] OR phospho-histon*[Title/Abstract] OR phosphohiston*[Title/Abstract] OR carcinogen*[Title/Abstract] OR "Carcinogens"[MeSH] OR "Carcinogenesis"[MeSH] OR mutagen*[Title/Abstract] OR "Mutagens"[MeSH] OR "Mutagenesis"[MeSH] OR mutation*[Title/Abstract] OR "Mutation"[MeSH] OR teratogen*[Title/Abstract] OR "Teratogens"[MeSH] OR "Teratogenesis"[MeSH] OR genetic*[Title/Abstract] OR "DNA"[Title/Abstract] OR "DNA"[Mesh] OR "RNA"[Title/Abstract] OR "RNA"[Mesh] OR "DNA Damage"[MeSH] OR insult*[Title/Abstract] OR adduct*[Title/Abstract] OR alkylation*[Title/Abstract] OR "Alkylation"[MeSH] OR alkylating[Title/Abstract] OR methylation*[Title/Abstract] OR "Methylation"[MeSH] OR oxidizing[Title/Abstract] OR oxidant*[Title/Abstract] OR "Oxidative Stress"[MeSH] OR "oxidative stress*"[Title/Abstract]) AND ("agrochemical*"[Title/Abstract] OR "agrichemical*"[Title/Abstract] OR "plant protection product*"[Title/Abstract] OR "pesticid*"[Title/Abstract] OR "biocid*"[Title/Abstract] OR "herbicid*"[Title/Abstract] OR "weedkiller*"[Title/Abstract] OR "weed killer*"[Title/Abstract] OR "defolian*"[Title/Abstract] OR "insecticid*"[Title/Abstract] OR "nematicid*"[Title/Abstract] OR "molluscicid*"[Title/Abstract] OR "piscicid*"[Title/Abstract] OR "avicid*"[Title/Abstract] OR "rodenticid*"[Title/Abstract] OR "repellent*"[Title/Abstract] OR "antiparasit*"[Title/Abstract] OR "lampricid*"[Title/Abstract] OR "acaricid*"[Title/Abstract] OR "miticid*"[Title/Abstract] OR "mite control*"[Title/Abstract] OR "algicid*"[Title/Abstract] OR "algaecid*"[Title/Abstract] OR "chemosterilant*"[Title/Abstract] OR "Agrochemicals"[MeSH:NoExp] OR "Pesticides"[MeSH])) AND ("Infant*"[Title/Abstract] OR "Prenatal*"[Title/Abstract] OR "postnatal*"[Title/Abstract] OR "antenatal*"[Title/Abstract] OR "Embryonic and Fetal Development"[Mesh] OR "Embryo*" [Title/Abstract] OR "Fetal*"[Title/Abstract] OR "foetal*"[Title/Abstract] OR "foetus*"[Title/Abstract] OR "fetus*"[Title/Abstract] OR "Gestational Age*"[Title/Abstract] OR "Infant"[Mesh] OR "Prenatal Diagnosis"[Mesh]) | 2,259 in 12/02/2024 |
| Web of science | "infant*" OR "Prenatal*" OR “postnatal*” OR “antenatal*” OR "embryo*" OR “fetal*” OR “foetal*” OR “foetus*” OR “fetus*” OR “Gestation*” (Topic) and “agrochemical*” OR “agrichemical*” OR "plant protection product*" OR “pesticid*” OR “biocid*” OR “herbicid*” OR “weedkiller*” OR "weed killer*" OR “defolian*” OR “insecticid*” OR “nematicid*” OR “molluscicid*” OR “piscicid*” OR “avicid*” OR “rodenticid*” OR “repellent*” OR "antiparasit*" OR “lampricid*” OR “acaricid*” OR “miticid*” OR "mite control*" OR “algicid*” OR “algaecid*” OR “chemosterilant*” (Topic) and "genotox*" OR "pharmacogenomic*" OR "comet assay*" OR "micronucleus*" OR "aberrat*" OR "chromosom*" OR "denaturation*" OR "hybridization*" OR "ames" OR "aneuploid*" OR "topoisomeras*" OR "teniposid*" OR "etoposid*" OR "greenscreen*" OR "γh2ax*" OR "ph2ax*" OR "high content screening*" OR "cycle arrest*" OR "phospho-histon*" OR "phosphohiston*" OR "caspase*" OR "tubulin microtubule*" OR "profiling assay*" OR "steatosis*" OR "carcino*" OR "mutagen*" OR "mutation*" OR "teratogen*" OR "genetic*" OR "dna" OR "rna" OR "damag*" OR "insult*" OR "adduct*" OR "alkylat*" OR "methylation*" OR "oxidiz*" OR "oxidant*" OR "oxidative stress*" OR "polymorph*" OR "gene tox*" OR "gene-tox*" OR "geno-tox*" (Topic) | 2,918 in 12/02/2024 |
| Scopus | **( TITLE-ABS-KEY ( "Infant*" OR "Prenatal*" OR "postnatal*" OR "antenatal*" OR "embryo*" OR "fetal*" OR "foetal*" OR "foetus*" OR "fetus*" OR "Gestation*" ) AND TITLE-ABS-KEY ( "agrochemical*" OR "agrichemical*" OR "plant protection product*" OR "pesticid*" OR "biocid*" OR "herbicid*" OR "weedkiller*" OR "weed killer*" OR "defolian*" OR "insecticid*" OR "nematicid*" OR "molluscicid*" OR "piscicid*" OR "avicid*" OR "rodenticid*" OR "repellent*" OR "antiparasit*" OR "lampricid*" OR "acaricid*" OR "miticid*" OR "mite control*" OR "algicid*" OR "algaecid*" OR "chemosterilant*" ) AND TITLE-ABS-KEY ("genotox*" OR "pharmacogenomic*" OR "comet assay*" OR "micronucleus*" OR "aberrat*" OR "chromosom*" OR "denaturation*" OR "hybridization*" OR "ames" OR "aneuploid*" OR "topoisomeras*" OR "teniposid*" OR "etoposid*" OR "greenscreen*" OR "γh2ax*" OR "ph2ax*" OR "high content screening*" OR "ph3" OR "cycle arrest*" OR "phospho-histon*" OR "phosphohiston*" OR "caspase*" OR "tubulin microtubule*" OR "profiling assay*" OR "steatosis*" OR "carcino*" OR "mutagen*" OR "mutation*" OR "teratogen*" OR "genetic*" OR "dna" OR "rna" OR "damag*" OR "insult*" OR "adduct*" OR "alkylat*" OR "methylation" OR "oxidizing" OR "oxidant*" OR "oxidative stress*"** OR "polymorph*" OR "gene tox*" OR "gene-tox*" OR "geno-tox*"**) )** | 5,417 in 12/02/2024 |
| Embase | **('infant*':ti,ab,kw OR 'prenatal*':ti,ab,kw OR 'postnatal*':ti,ab,kw OR 'antenatal*':ti,ab,kw OR 'embryo*':ti,ab,kw OR 'fetal*':ti,ab,kw OR 'foetal*':ti,ab,kw OR 'foetus*':ti,ab,kw OR 'fetus*':ti,ab,kw OR 'gestation*':ti,ab,kw) AND ('agrochemical*':ti,ab,kw OR 'agrichemical*':ti,ab,kw OR 'plant protection product*':ti,ab,kw OR 'pesticid*':ti,ab,kw OR 'biocid*':ti,ab,kw OR 'herbicid*':ti,ab,kw OR 'weedkiller*':ti,ab,kw OR 'weed killer*':ti,ab,kw OR 'defolian*':ti,ab,kw OR 'insecticid*':ti,ab,kw OR 'nematicid*':ti,ab,kw OR 'molluscicid*':ti,ab,kw OR 'piscicid*':ti,ab,kw OR 'avicid*':ti,ab,kw OR 'rodenticid*':ti,ab,kw OR 'repellent*':ti,ab,kw OR 'antiparasit*':ti,ab,kw OR 'lampricid*':ti,ab,kw OR 'acaricid*':ti,ab,kw OR 'miticid*':ti,ab,kw OR 'mite control*':ti,ab,kw OR 'algicid*':ti,ab,kw OR 'algaecid*':ti,ab,kw OR 'chemosterilant*':ti,ab,kw) AND ('**genotox***':ti,ab,kw** OR **'**pharmacogenomic***':ti,ab,kw** OR **'**comet assay***':ti,ab,kw** OR **'**micronucleus***':ti,ab,kw** OR **'**aberrat***':ti,ab,kw** OR **'**chromosom***':ti,ab,kw** OR **'**denaturation***':ti,ab,kw** OR **'**hybridization***':ti,ab,kw** OR **'**ames**':ti,ab,kw** OR **'**aneuploid***':ti,ab,kw** OR **'**topoisomeras***':ti,ab,kw** OR **'**teniposid***':ti,ab,kw** OR **'**etoposid***':ti,ab,kw** OR **'**greenscreen***':ti,ab,kw** OR **'**γh2ax***':ti,ab,kw** OR **'**ph2ax***':ti,ab,kw** OR **'**high content screening***':ti,ab,kw** OR **'**ph3**':ti,ab,kw** OR "cycle arrest***':ti,ab,kw** OR **'**phospho-histon***':ti,ab,kw** OR **'**phosphohiston***':ti,ab,kw** OR **'**caspase***':ti,ab,kw** OR **'**tubulin microtubule***':ti,ab,kw** OR **'**profiling assay***':ti,ab,kw** OR **'**steatosis***':ti,ab,kw** OR **'**carcino***':ti,ab,kw** OR **'**mutagen***':ti,ab,kw** OR **'**mutation***':ti,ab,kw** OR **'**teratogen***':ti,ab,kw** OR **'**genetic***':ti,ab,kw** OR **'**dna**':ti,ab,kw** OR **'**rna**':ti,ab,kw** OR **'**damag***':ti,ab,kw** OR **'**insult***':ti,ab,kw** OR **'**adduct***':ti,ab,kw** OR **'**alkylat***':ti,ab,kw** OR **'**methylation***':ti,ab,kw** OR **'**oxidizing**':ti,ab,kw** OR **'**oxidant***':ti,ab,kw** OR **'**oxidative stress***':ti,ab,kw** OR **'**polymorph***':ti,ab,kw** OR **'**gene tox***':ti,ab,kw** OR **'**gene-tox***':ti,ab,kw** OR **'**geno-tox***':ti,ab,kw)** | 2,190 in 12/02/2024 |
